# Factors associated with tuberculosis treatment initiation among bacteriologically negative individuals evaluated for tuberculosis: an individual patient data meta-analysis

**DOI:** 10.1101/2024.04.07.24305445

**Authors:** Sun Kim, Melike Hazal Can, Tefera B. Agizew, Andrew F. Auld, Maria Elvira Balcells, Stephanie Bjerrum, Keertan Dheda, Susan E. Dorman, Aliasgar Esmail, Katherine Fielding, Alberto L. Garcia-Basteiro, Colleen F. Hanrahan, Wakjira Kebede, Mikashmi Kohli, Anne F. Luetkemeyer, Carol Mita, Byron W. P. Reeve, Denise Rossato Silva, Sedona Sweeney, Grant Theron, Anete Trajman, Anna Vassall, Joshua L. Warren, Marcel Yotebieng, Ted Cohen, Nicolas A. Menzies

## Abstract

**Background:** Globally, over one-third of pulmonary tuberculosis (TB) disease diagnoses are made based on clinical criteria after a negative diagnostic test result. Understanding factors associated with clinicians’ decisions to initiate treatment for individuals with negative test results is critical for predicting the potential impact of new diagnostics.

**Methods:** We performed a systematic review and individual patient data meta-analysis using studies conducted between January/2010 and December/2022 (PROSPERO: CRD42022287613). We included trials or cohort studies that enrolled individuals evaluated for TB in routine settings. In these studies participants were evaluated based on clinical examination and routinely-used diagnostics, and were followed for ≥1 week after the initial test result. We used hierarchical Bayesian logistic regression to identify factors associated with treatment initiation following a negative result on an initial bacteriological test (e.g., sputum smear microscopy, Xpert MTB/RIF).

**Findings:** Multiple factors were positively associated with treatment initiation: male sex [adjusted Odds Ratio (aOR) 1.61 (1.31–1.95)], history of prior TB [aOR 1.36 (1.06–1.73)], reported cough [aOR 4.62 (3.42–6.27)], reported night sweats [aOR 1.50 (1.21–1.90)], and having HIV infection but not on ART [aOR 1.68 (1.23–2.32)]. Treatment initiation was substantially less likely for individuals testing negative with Xpert [aOR 0.77 (0.62–0.96)] compared to smear microscopy and declined in more recent years.

**Interpretation:** Multiple factors influenced decisions to initiate TB treatment despite negative test results. Clinicians were substantially less likely to treat in the absence of a positive test result when using more sensitive, PCR-based diagnostics.

**Funding:** National Institutes of Health

**Research in context:** *Evidence before this study:* In countries with a high burden of tuberculosis, over one-third of notified cases for pulmonary TB are diagnosed based on clinical criteria, without bacteriological confirmation of disease (‘clinical diagnosis’). For these individuals with negative bacteriological test results, there is limited evidence on the factors associated with higher or lower rates of clinical diagnosis. In the context of individual clinical trials, some analyses have reported lower rates of treatment initiation for individuals testing negative on new cartridge-based PCR tests (e.g., Xpert MTB-RIF), as compared to individuals testing negative in sputum smear microscopy.

*Added value of this study:* This study conducted a systematic review of studies that collected data on patient characteristics and treatment initiation decisions for individuals receiving a negative bacteriological test result as part of initial evaluation for TB. Patient-level data from 13 countries across 12 studies (n=15121) were analyzed in an individual patient data meta-analysis, to describe factors associated with clinicians’ decisions to treat for TB disease. We identified significant associations between multiple clinical factors and the probability that a patient would be initiated on TB treatment, including sex, history of prior TB, reported symptoms (cough and night sweats), and HIV status. Controlling for other factors, patients testing negative on PCR-based diagnostics (e.g., Xpert MTB/RIF) were less likely to be initiated on treatment than those testing negative with smear microscopy.

*Implications of all the available evidence:* Rates of clinical diagnosis for TB differ systematically as a function of multiple clinical factors and are lower for patients who test negative with new PCR-based diagnostics compared to earlier smear-based methods. This evidence can be used to refine diagnostic algorithms and better understand the implications of introducing new diagnostic tests for TB.

## INTRODUCTION

Tuberculosis (TB) remains a leading cause of infectious disease death worldwide (1), and a key strategy for accelerating TB elimination is to improve capacity for rapid and accurate diagnosis in high burden countries (2). Traditional TB diagnostics have major limitations, with sputum smear microscopy (SSM) failing to identify a substantial fraction of TB cases, and sputum culture requiring up to eight weeks to return results. However, since 2010 the World Health Organization (WHO) has endorsed several new PCR (polymerase chain reaction)-based diagnostics with the potential to improve TB case detection, including the Xpert MTB/RIF (Xpert), Xpert MTB/RIF Ultra (Xpert Ultra), Truenat MTB, Truenat MTB Plus, and Truenat MTB-RIF Dx assays (3). Compared to SSM and culture, these tests combine rapid turn-around time and high sensitivity, enabling timely and accurate TB diagnosis (3,4).

Despite the potential of these new diagnostics, several studies have found limited effects on TB diagnoses and mortality following their introduction (5–13). Evidence from programmatic settings suggests that clinical diagnosis (diagnosis based on clinical criteria alone, made when a bacteriological test result is unavailable or is negative) may partially explain this finding (14–17). In many countries, clinical diagnosis represents a substantial fraction of notified TB cases despite the widespread adoption of Xpert, and in 2022 clinical diagnoses represented 38% of total global notifications for pulmonary TB (1). If some of the individuals testing false-negative on an initial bacteriological test are subsequently treated based on clinical criteria, this may reduce the incremental impact achieved by adopting a more sensitive diagnostic. However, the widespread use of clinical diagnosis may also increase the number of individuals incorrectly treated for TB and overlook cases of drug-resistant TB, as studies of the performance of clinical diagnosis suggest that the specificity of clinical algorithms can be low (18–20). For certain types of tuberculosis, such as extrapulmonary and pediatric TB, clinical diagnosis may be the sole viable option.

As higher sensitivity diagnostics become more commonly used it is useful to understand current practices around clinical diagnosis, and the factors that affect clinical decision-making when diagnostic test results are negative. These clinical decisions will affect the overall sensitivity and specificity of TB diagnostic algorithms, as well as determining the incremental health impact of new diagnostics. In this study, we conducted a systematic review of studies reporting diagnostic decisions and treatment initiation following a negative test result received as part of routine TB diagnosis. Using these data, we conducted an individual patient data (IPD) meta-analysis to identify the factors that affect clinicians’ decisions to treat for pulmonary TB despite a negative test result.

## METHODS

The target population for this study was individuals evaluated for pulmonary TB disease in routine clinical settings, who had received a negative result on an initial diagnostic test (e.g., smear microscopy, Xpert MTB/RIF). We conducted a systematic review to identify datasets describing the individual characteristics as well as the outcome of TB diagnosis (i.e., whether or not TB treatment was initiated) for individuals in this target population. The protocol was registered with PROSPERO (CRD42022287613) and approved by the Institutional Review Board of the Harvard School of Public Health (IRB21-1488).

### Search strategy and selection criteria

Studies were identified by searching Medline / PubMed and Embase (search strategies for each database provided in Supplement). The publication date was limited to 2010 - 2022 in order to restrict the analysis to the period over which new TB diagnostics were being introduced. We also contacted subject matter experts to identify ongoing or recently completed studies not identified in the database search.

Studies eligible for the review included randomized controlled studies or cohort studies (a) that enrolled individuals evaluated for TB after presenting for care at routine healthcare settings, (b) where treatment decisions were based on diagnostic tests in routine use in that setting (i.e., additional tests conducted for research purposes were not used), and (c) where participants were followed for least 1 week following the initial diagnostic test to record whether or not treatment was initiated. We excluded systematic reviews and studies of non-human subjects, pediatric TB, latent TB, hospitalized patients (in-patient), multi-drug resistant TB, and active case finding. De-identified patient-level data were obtained by contacting the investigators of studies meeting inclusion criteria.

### Variables of interest

For each study dataset, we extracted data on individual-level variables describing the type of initial test received (e.g., Xpert, Ultra, Truenat, SSM), age (18 years or older), sex, presence of TB-related symptoms (cough, fever, night sweats, weight loss), results for any non-bacteriological tests performed (e.g., chest radiography), HIV status, morbidity score (e.g., Karnofsky score ranging from 0 (dead) to 100 indicating no evidence of disease) if available, TB diagnosis, whether TB treatment was initiated, date of treatment initiation, date of testing, date culture result was returned (if applicable), and duration of follow-up. We also extracted contextual variables including calendar year, country, and type of clinic at which the patient was evaluated (primary, secondary). We excluded individuals with inconclusive or missing results for the initial diagnostic test.

After data extraction we created a master list of variables available from each study. Relevant variables that could influence diagnostic decision-making were selected based on TB diagnostic algorithms and guidelines consolidated by WHO (3,21). Given that each study has different variables and units, we selected common variables across studies for meta-analysis and converted variable types for consistency across studies (e.g., conversion of continuous variables to categorical variables for symptom durations (unit in weeks)). We collated the harmonized individual patient data (IPD) into a single dataset.

Our primary outcome was whether or not an individual initiated TB treatment following a negative SSM, Xpert, or Xpert Ultra result (i.e., the standard of care for initial TB testing in each setting at the time of the study). While some studies undertook additional investigational tests clinicians were blinded to these results. Although most studies collected samples for sputum culture, we restricted our analysis to the period before culture results became available.

For studies that recorded a variable indicating whether or treatment was provided on clinical grounds, we used this variable as our outcome measure. For all other studies we defined clinical diagnosis as instances where treatment was initiated following negative initial test results but before culture results became available.

### Data analysis

IPD meta-analysis was performed via logistic regression, specified for the binary outcome of whether or not an individual initiated treatment as defined above. To do so, we employed a hierarchical Bayesian model with country random effects (see Supplement), to account for country-specific differences in diagnostic practices not reflected in other variables (22,23). For the primary analysis we fit univariable and multivariable regression models considering age (18– 30 years, 31–40 years, >40 years), sex (female, male), history of prior TB (no, yes, unknown), reported cough (yes, no), reported night sweats (yes, no), reported fever (yes, no), HIV status (negative, positive (not on ART), positive (on ART), unknown), test type (SSM, Xpert, Xpert Ultra), and calendar year. These variables were included in the primary analysis based on their availability in the majority of datasets.

We conducted two secondary analyses using variables not available for a subset of datasets. First, using the datasets that provided information on symptom duration, we fit a modified version of the regression model for the primary analysis, in which the binary variables for cough, fever, and night sweats were replaced by versions of these variables that each stratified the observations into one of three levels (none, less than two weeks, two weeks and above).

Second, for the datasets containing chest x-ray results, we reran the regression model for the primary analysis with this additional variable (normal, abnormal, unknown).

As a robustness check we re-estimated the results of the main analysis with two alternative regression specifications. First, we adopted an alternative outcome definition, in which clinical diagnosis was defined as treatment initiation within 7 days of the initial diagnostic test. While potentially excluding some clinical diagnoses, this stricter definition may reduce the risk of bias due to variation in the definition of clinical diagnosis adopted by each study. Second, we re-estimated results using a regression model in which the country random effects were replaced by study random effects. All statistical analyses were performed in R (v.4.2.3) using the “brms” package (v.2.19.0) (24–26).

## RESULTS

Our database search identified 4,286 potentially eligible studies. After removal of duplicates this resulted in 3,428 unique references for screening. After review of title and abstract of those references, full-text screening was performed on 161 studies, with 51 eligible studies identified (**Figure 1**). Following communication with investigators for each study we obtained data from 18 eligible studies. Six of these studies were excluded after initial data cleaning due to missing key variables or considering a different target population. The final dataset included observations collected between 2011 and 2020, covering 13 countries across 12 studies. Most of these countries are classified as high-burden for TB by the WHO. **Table 1** reports demographic and clinical characteristics for the full analytic sample, and **Table S1** provides details of each included study.

**Figure 1.**
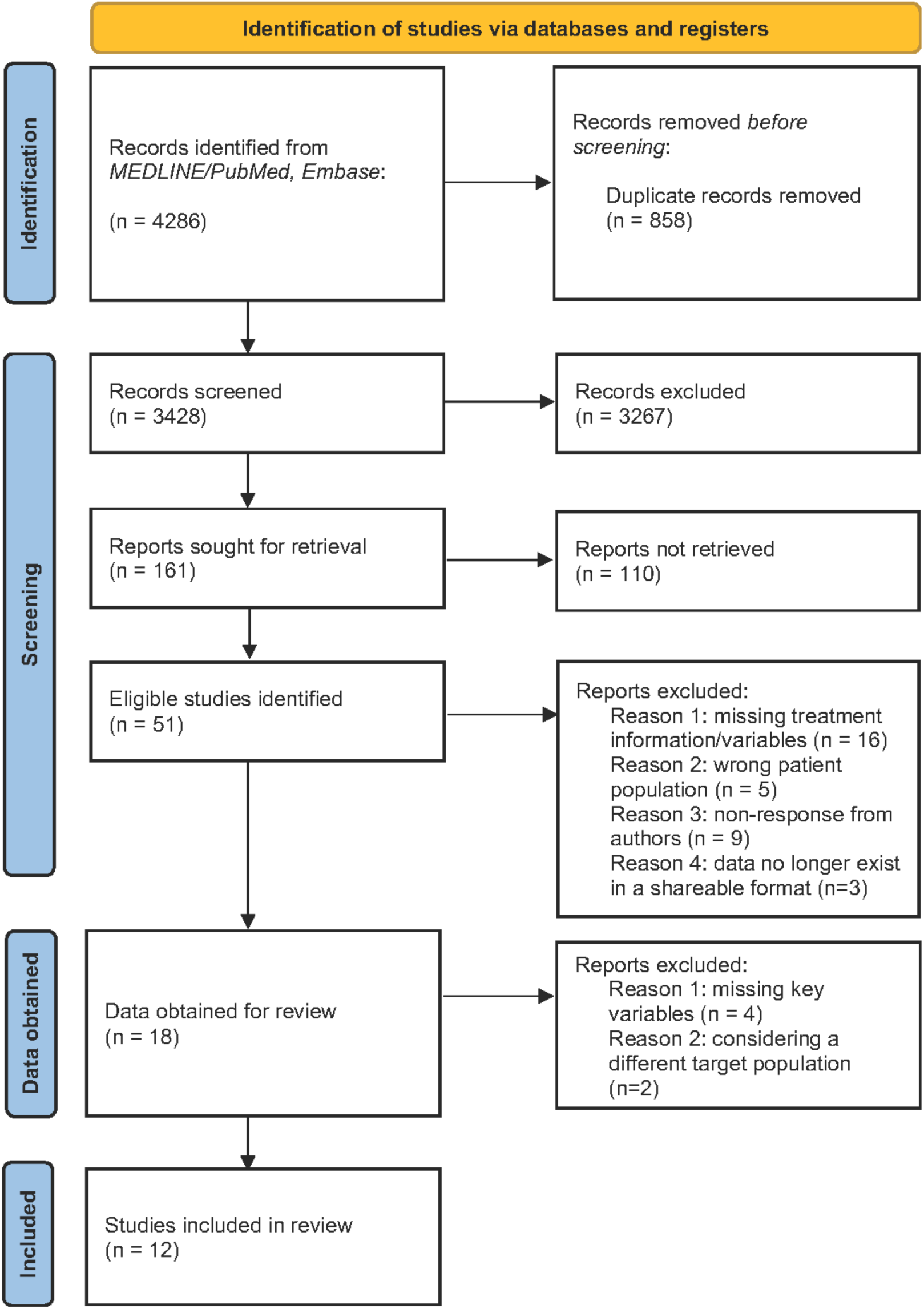
Identification of studies and data to include in the meta-analysis.

**Table 1.**
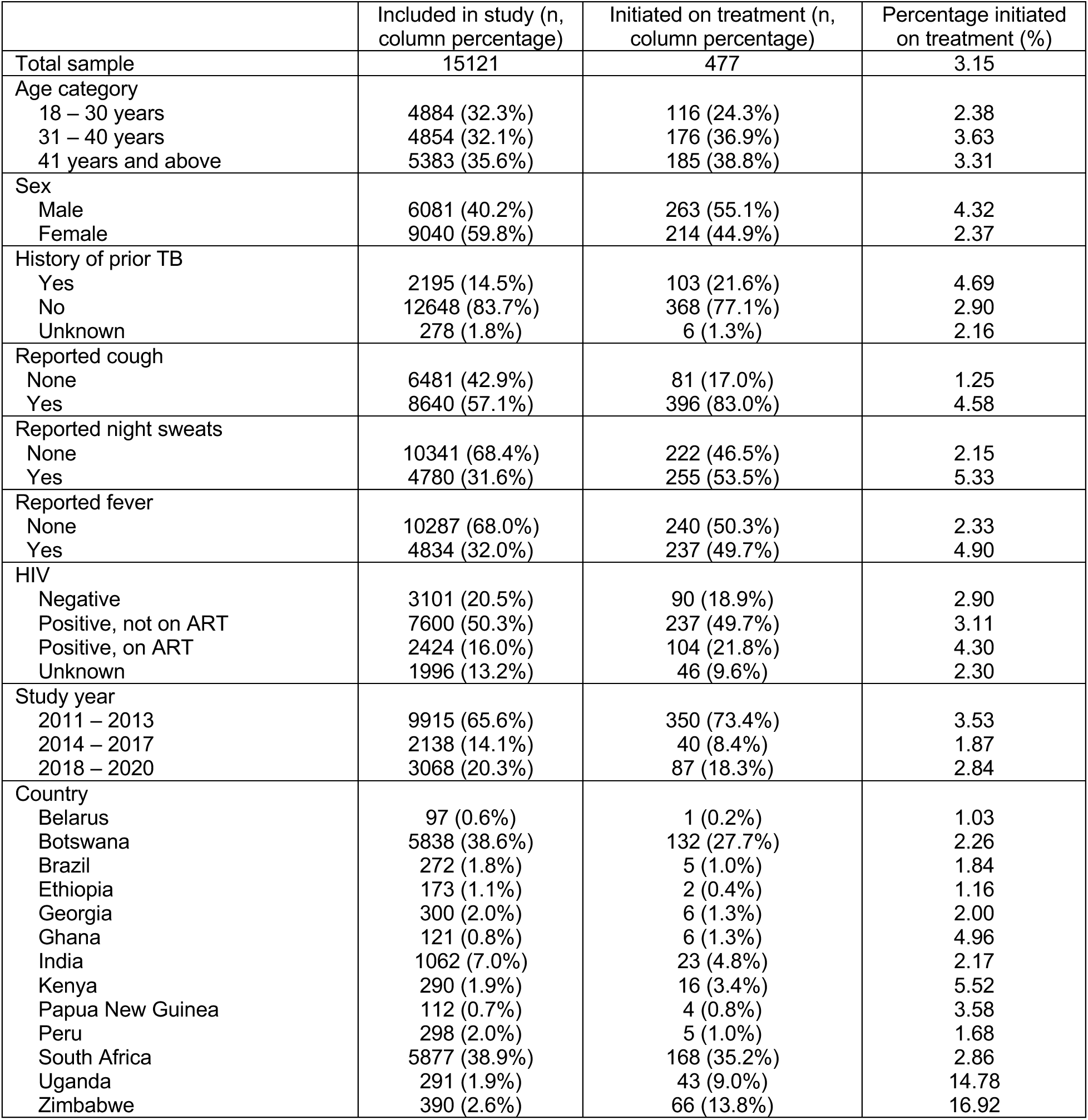
Demographic and clinical characteristics of study population.

### Primary analysis

The main analysis included data for 15,121 adults evaluated for pulmonary TB for whom the initial TB test was negative. Of these individuals 477 were initiated on TB treatment following clinical diagnosis. **Table 2** summarizes the meta-analysis results as odds ratios (OR) and adjusted odds ratios (aOR) produced by univariable and multivariable regression models respectively, representing the odd ratio of TB treatment initiation among individuals with a given factor compared to the reference category.

**Table 2.** Odds ratios of TB treatment initiation following a negative diagnostic test result. *Multivariable regression model also included country random effects, coefficients shown in Table S2. Ref = reference category.

|  | Univariable analysis<br>(95% Credible Intervals) | Multivariable analysis*<br>(95% Credible Intervals) |
| --- | --- | --- |
| Age category |  |  |
| 18 – 30 years | Ref | Ref |
| 31 – 40 years | 1.44 (1.13–1.83) | 1.17 (0.92–1.51) |
| 41 years and above | 1.42 (1.12–1.80) | 1.11 (0.87–1.43) |
| Sex |  |  |
| Female | Ref | Ref |
| Male | 1.80 (1.50–2.16) | 1.61 (1.31–1.95) |
| History of prior TB |  |  |
| None | Ref | Ref |
| Yes | 1.61 (1.13–2.03) | 1.36 (1.06–1.73) |
| Unknown | 0.81 (0.32–1.77) | 0.73 (0.28–1.65) |
| Reported cough |  |  |
| None | Ref | Ref |
| Yes | 5.93 (4.52–7.88) | 4.62 (3.42–6.27) |
| Reported night sweats |  |  |
| None | Ref | Ref |
| Yes | 2.34 (1.89–2.89) | 1.50 (1.21–1.90) |
| Reported fever |  |  |
| None | Ref | Ref |
| Yes | 1.84 (1.48–2.28) | 1.13 (0.91–1.39) |
| HIV |  |  |
| Negative | Ref | Ref |
| Positive, not on ART | 1.88 (1.38–2.57) | 1.68 (1.23–2.32) |
| Positive, on ART | 0.73 (0.52–1.02) | 0.90 (0.64–1.30) |
| Unknown | 0.98 (0.65–1.46) | 0.82 (0.55–1.20) |
| Diagnostic test |  |  |
| Sputum Smear | Ref | Ref |
| Xpert | 0.64 (0.51–0.79) | 0.77 (0.62–0.96) |
| Xpert Ultra | 0.21 (0.13–0.34) | 0.57 (0.30–1.07) |
| Year | 0.77 (0.71–0.83) | 0.81 (0.74–0.90) |

Based on the multivariable analysis, we identified statistically significant increases in the odds of TB treatment initiation associated with male sex (aOR 1.61 compared to female sex, 95% credible interval (CI): 1.31–1.95), having a history of prior TB (aOR 1.36 compared to individuals without prior TB, 95% CI: 1.06–1.73), having reported cough (aOR 4.62 compared to no cough, 95% CI: 3.42–6.27), having reported night sweats (aOR 1.50 compared to no night sweats, 95% CI: 1.21–1.90), and having HIV infection but not on ART (aOR 1.68 compared to HIV-negative, 95% CI: 1.23–2.32).

In terms of the tests used for initial TB diagnosis, we found lower odds of treatment initiation for individuals who had received a negative result on Xpert (aOR 0.77 compared to diagnosis via SSM, 95% CI: 0.62–0.96) and who had received a negative result on Xpert Ultra (aOR 0.57 compared to diagnosis via SSM, 95% CI: 0.30–1.07), although the results for Xpert Ultra were not statistically significant. We also estimated declining rates of treatment initiation over time, controlling for other factors (aOR 0.81 for each additional calendar year, 95% CI: 0.74–0.90).

### Secondary analyses

In the first secondary analysis, we estimated odds ratios for cough, fever, and night sweats categorized by duration of symptoms, using data from the five studies for which this variable was available (7,468 observations). These findings indicated strong positive associations between TB treatment initiation and a reported cough of 0-2 weeks duration (aOR 3.29 compared to no reported cough, 95% CI: 1.64–7.34) and >2 weeks duration (aOR 5.34 compared to no reported cough, 95% CI: 2.72–11.82) (**Figure 2**). Reported night sweats of 0-2 weeks duration also demonstrated elevated odds of treatment initiation (aOR 1.45 compared to no reported night sweats, 95% CI: 1.06–2.00).

**Figure 2.**
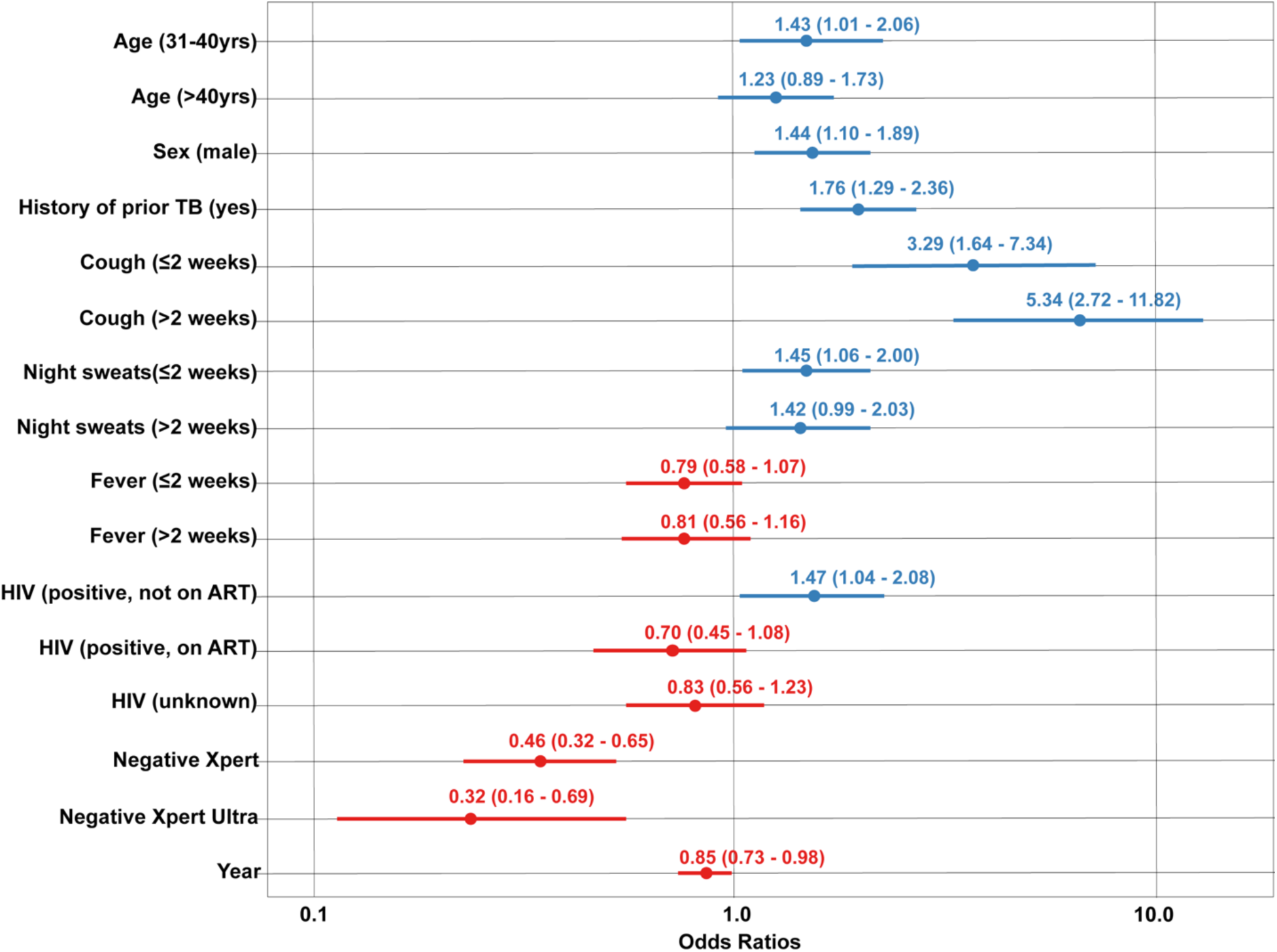
Odds ratios of TB treatment initiation following negative diagnostic test result: secondary analysis for datasets including duration of symptoms for cough, fever, and night sweats*. * Reference group: Age 18-30 years old, female sex, no history of prior TB, no reported cough, no reported fever, no reported night sweats, HIV-negative, tested negative with sputum smear microscopy. Blue symbols signify odds ratios >1.0, red symbols signify odds ratios <1.0.

The second secondary analysis estimated differences in treatment initiation based on chest x-ray result, using data from the three studies in which x-ray was conducted as part of TB evaluation (2,449 observations). In these data 1,677 individuals had a normal x-ray result (1.1% (18/1677) initiated on treatment) and 456 had an abnormal x-ray result (6.1% (28/456) initiated on treatment). The results of this analysis showed that having an abnormal X-ray result had a strong positive association with treatment initiation, with an adjusted odds ratio of 6.89 (95% CI: 3.29–14.42) compared to individuals with normal x-ray results (**Table S3**).

### Alternative model specifications

**Table 3** presents results for two alternative model specifications. In the first alternative specification we analyzed an alternative outcome defined as treatment initiation within 7 days of the initial diagnostic test, representing 1.4% (205/15121) of all observations. These results were generally consistent with the results of the primary analysis, although the odds ratio estimated for receiving a negative result on Xpert Ultra was lower than in the primary analysis and statistically significant (aOR 0.35 compared to diagnosis via SSM, 95% CI: 0.17–0.75). Additionally, the estimated time trend in treatment initiation was no longer significant (aOR 0.97 for each additional calendar year, 95% CI: 0.85–1.09).

**Table 3.** Odds ratios of TB treatment initiation following negative diagnostic test result: alternative model specification using strict outcome definition and study random effects. *Our primary outcome was the initiation of TB treatment after negative SSM, Xpert, or Xpert Ultra results; using treatment provision on clinical grounds if available, otherwise defining clinical diagnosis as treatment initiation post-negative initial tests prior to culture results. **For alternative outcome definition, clinical diagnosis was defined as treatment initiation within 7 days of the initial diagnostic test. ***Multivariable regression model also included study random effects, coefficients shown in **Table S4**. Ref = reference category.

|  | Adjusted odds ratios for treatment initiation (95% Credible Intervals) |  |  |
| --- | --- | --- | --- |
|  | Main analysis<br>(primary outcome*,<br>country random effects) | First alternative<br>specification<br>(alternative outcome**,<br>country random effects) | Second alternative<br>specification<br>(primary outcome, study<br>random effects)*** |
| Age category |  |  |  |
| 18 – 30 years | Ref | Ref | Ref |
| 31 – 40 years | 1.17 (0.92–1.51) | 1.35 (0.90–2.02) | 1.13 (0.88–1.46) |
| 41 years and above | 1.11 (0.87–1.43) | 1.37 (0.94–2.06) | 1.07 (0.84–1.37) |
| Sex |  |  |  |
| Female | Ref | Ref | Ref |
| Male | 1.61 (1.31–1.95) | 1.57 (1.17–2.12) | 1.62 (1.34–1.95) |
| History of prior TB |  |  |  |
| None | Ref | Ref | Ref |
| Yes | 1.36 (1.06–1.73) | 1.45 (1.03–2.02) | 1.36 (1.08–1.72) |
| Unknown | 0.73 (0.28–1.65) | 0.57 (0.14–1.75) | 0.39 (0.06–1.65) |
| Reported cough |  |  |  |
| None | Ref | Ref | Ref |
| Yes | 4.62 (3.42–6.27) | 6.78 (3.78–12.65) | 4.73 (3.50–6.37) |
| Reported night sweats |  |  |  |
| None | Ref | Ref | Ref |
| Yes | 1.50 (1.21–1.90) | 1.29 (0.93–1.78) | 1.45 (1.17–1.82) |
| Reported fever |  |  |  |
| None | Ref | Ref | Ref |
| Yes | 1.13 (0.91–1.39) | 1.14 (0.84–1.56) | 1.08 (0.87–1.34) |
| HIV |  |  |  |
| Negative | Ref | Ref | Ref |
| Positive, not on ART | 1.68 (1.23–2.32) | 1.84 (1.16–2.95) | 1.55 (1.12–2.14) |
| Positive, on ART | 0.90 (0.64–1.30) | 0.92 (0.58–1.48) | 0.86 (0.59–1.24) |
| Unknown | 0.82 (0.55–1.20) | 0.38 (0.20–0.69) | 0.87 (0.59–1.29) |
| Diagnostic test |  |  |  |
| Sputum Smear | Ref | Ref | Ref |
| Xpert | 0.77 (0.62–0.96) | 0.65 (0.42–0.98) | 0.79 (0.62–0.99) |
| Xpert Ultra | 0.57 (0.30–1.07) | 0.35 (0.17–0.75) | 0.37 (0.21–0.64) |
| Year | 0.81 (0.74–0.90) | 0.97 (0.85–1.09) | 0.87 (0.74–1.04) |

The results for the second alternative specification (results estimated with study random effects instead of country random effects) were generally consistent with the results of the primary analysis, although the odds ratio estimated for receiving a negative result on Xpert Ultra was lower than in the primary analysis and statistically significant (aOR 0.37 compared to diagnosis via SSM, 95% CI: 0.21–0.64). The estimated time trend in treatment initiation was no longer significant (aOR 0.87 for each additional calendar year, 95% CI: 0.74–1.04).

## DISCUSSION

This study examined the factors associated with treatment initiation among adults evaluated for TB in routine healthcare settings, who had received a negative result on an initial bacteriological test for TB. Our analyses showed that male sex, a history of prior TB, reported cough, and having HIV infection but not receiving ART were positively associated with clinicians’ decisions to initiate TB treatment. Among the three tests used for initial diagnosis, individuals receiving a negative result on Xpert were substantially less likely to be initiated on treatment compared to individuals who had received a negative result with SSM. Though not statistically significant in the main analysis, a negative result on Xpert Ultra was also associated with lower treatment initiation rates compared to SSM. In addition, the secondary analyses demonstrated increasing odds of treatment initiation with longer duration of cough (specifically, cough persisting for over two weeks). Similarly, the presence of an abnormal chest X-ray result was found to have a strong positive association with treatment initiation. We also observed a lower likelihood of treatment initiation in more recent years, controlling for other factors.

Most results from the alternative model specifications were consistent with those of the primary analysis. For the first alternative specification (outcome defined as treatment initiation within 7 days of the initial TB test) the fraction diagnosed clinically was lower than in the primary analysis (3.2% vs. 1.4%), and this outcome definition may have excluded some individuals who were treated clinically but with a greater delay. However, this outcome definition reduced potential inter-study variation in the definition of clinical diagnosis, and the risk of bias due to clinicians accessing culture results before making treatment decisions. The second alternative specification assumed that residual variation in clinical decision-making was primarily attributable to study-specific factors (vs. country-specific factors in the main analysis). That the estimated odds ratios were mostly consistent across different model specifications provides some assurance that these results are robust. One small difference was for Xpert Ultra, for which in both alternative specifications individuals testing negative on Xpert Ultra were estimated to be significantly less likely to begin treatment compared to those who received a negative result from SSM, with these odds ratios lower than those estimated in the primary analysis, and statistically significant. In addition, the results describing the time trend were no longer statistically significant in both alternative specifications.

The findings for individual covariates can be interpreted in light of factors that clinicians may consider during TB diagnosis. These considerations include the pre-test probability of disease (prevalence of TB disease among individuals being tested), the expected magnitude of harms resulting from an incorrect negative diagnosis relative to the harms of an incorrect positive diagnosis, and the expected sensitivity and specificity of the tests being used. Several of the covariates examined in this study are relevant to these considerations.

First, several of the covariates we examined may influence clinician’s beliefs about the pre-test probability of disease. Based on WHO guidelines for TB diagnosis and treatment in HIV-prevalent and resource constrained settings, a history of prior TB and symptoms suggestive of TB imply a higher pre-test probability of disease, and therefore may increase clinical suspicion for TB (27). Similarly, in many settings persons living with HIV have higher TB incidence compared to HIV-negative individuals, and men have elevated incidence rates compared to women, such that clinicians may expect these characteristics to imply a higher disease prevalence among those evaluated for TB. In light of these relationships (each of which was linked to elevated treatment initiation rates), it is somewhat surprising that reported fever had a modest association with treatment initiation. While the presence of fever has been associated with TB, it is also associated with many other conditions, and therefore may be of limited value in distinguishing TB from other alternative diagnoses (as has been found with antibiotic trial as a diagnostic modality (28)).

For the second consideration (harms resulting from an incorrect negative diagnosis relative to the harms from an incorrect positive diagnosis), it is possible that this contributes to the elevated treatment initiation odds estimated for persons living with HIV, as compared to HIV-negative individuals. Individuals with both HIV and TB experience rapid disease progression and are less likely to survive the TB episode compared to HIV-negative individuals with TB (29–31). As a consequence, the urgency of initiating TB treatment (if TB is suspected) will be much greater for individuals found to have HIV compared to those living without HIV. In contrast, the harms produced by a false-positive diagnosis, while not trivial, may not differ substantially between individuals with and without HIV.

For the third consideration (test sensitivity and specificity), this may explain the results estimated for the different test types (smear microscopy, Xpert, Xpert Ultra). The poor sensitivity of smear microscopy for pulmonary TB is well known, as is the improved performance of Xpert and Xpert Ultra compared to smear microscopy (32,33). Because of the higher sensitivity of these new PCR-based tests, an individual testing negative on one of these tests is less likely to have TB than if the individual had instead tested negative with smear, all other things being equal. Clinicians aware of these relationships may be more hesitant to recommend treatment for patients that have tested negative with a high-sensitivity test. It is also true that each of the tests examined is known to have lower sensitivity among individuals with HIV infection (34), and this may be an additional factor contributing to the higher odds of treatment initiation for HIV-positive individuals following a negative test.

There are several limitations to this study. First, we were not able to analyze all factors that potentially inform clinician decision making, due to differences in the covariates recorded in the study datasets. It is possible that additional individual characteristics—such as recent weight loss or reporting a known TB contact—may impact clinical decision-making but were not consistently captured in the study data. Similarly, it is possible that factors related to the healthcare setting or the clinicians performing diagnosis may influence rates of clinical diagnosis but were not available for analysis. Second, our analytic population excluded patients aged under 18. While diagnosis for older children and adolescents may be similar to adults, clinicians will have different decision criteria for diagnosis of infants, due to both the different presentation of TB and the poor performance of available TB diagnostics in young children. Third, while we selected studies to only include those performed under routine clinical conditions, it is possible that the behavior of clinicians performing TB diagnosis could have been influenced by their participation in clinical research. It is also possible that the clinics in which these studies were conducted were selected in a way that limits their representativeness of the general context of TB care. Fourth, while many of the findings of the analysis are consistent with general principles of good patient care (as discussed above), we did not have access to additional evidence describing why clinicians made the decisions they did. Fifth, we did not compare clinical diagnosis decisions with culture results that subsequently became available. While such a comparison was outside the scope of the current study—which focused on clinical decisions made before any additional test results became available—this comparison would be useful for judging the diagnostic accuracy of clinical diagnosis, and could be addressed in a subsequent study.

In conclusion, in this multi-country IPD meta-analysis of clinical diagnosis for TB, we found multiple clinical factors to be associated with the decision to initiate TB among individuals who receive a negative result on an initial bacteriological test for TB. Understanding these factors will allow for a more nuanced interpretation of the data describing the impact of introducing new TB diagnostics (35–37), and can inform efforts to refine clinical diagnostic algorithms, determine the appropriate balance between sensitivity and specificity when revising diagnostic approaches (38), and improve the overall performance of TB case detection.

## Data Availability

All data produced in the present study are available upon reasonable request to the authors.

