## Supplement for "Factors associated with tuberculosis treatment initiation among bacteriologically negative individuals evaluated for tuberculosis: an individual patient data meta-analysis"

| | Churchyard et al., 2015 | Penn-Nicholson et al., 2021 | Dorman et al., unpublished | Dorman et al., 2018 | Theron et al., unpublished | Mupfumi et al., 2014 |
|---|---|---|---|---|---|---|
| Study sites eligible for inclusion* | South Africa | Ethiopia, India, Papua New Guinea, and Peru | Kenya, South Africa, and Uganda | Belarus, Georgia, India, and South Africa | South Africa | Zimbabwe |
| Analytic population, n | 4001 | 1411 | 906 | 870 | 777 | 390 |
| Individuals with treatment initiation, n (primary outcome) | 130 | 22 | 61 | 23 | 4 | 66 |
| Alternative outcome, n | 20 | 19 | 61 | 23 | 1 | 16 |
| Age category, n | | | | | | |
| 18 – 30 years | 1322 | 370 | 263 | 206 | 340 | 80 |
| 31 – 40 years | 1134 | 275 | 282 | 214 | 267 | 174 |
| 41 years and above | 1545 | 766 | 361 | 450 | 170 | 136 |
| Male, n | 1444 | 784 | 459 | 522 | 300 | 171 |
| Individuals with history of prior TB, n | 613 | 192 | 223 | 255 | 107 | 49 |
| Reported cough | | | | | | |
| None | 759 | 0 | 0 | 48 | 566 | 200 |
| Yes | 3242 | 1411 | 906 | 822 | 211 | 190 |
| Reported night sweats | | | | | | |
| None | 2254 | 829 | 310 | 433 | 618 | 188 |
| Yes | 1747 | 582 | 596 | 437 | 159 | 202 |
| Reported fever | | | | | | |
| None | 2219 | 629 | 275 | 360 | 742 | 161 |
| Yes | 1782 | 782 | 631 | 510 | 35 | 229 |
| HIV | | | | | | |
| Negative | 1203 | 703 | 429 | 318 | 0 | 0 |
| Positive, not on ART | 1246 | 0 | 64 | 100 | 0 | 61 |
| Positive, on ART | 624 | 28 | 412 | 67 | 777 | 329 |
| Unknown | 928 | 680 | 1 | 385 | 0 | 0 |
| Individuals with abnormal chest X-ray result, n | NA | 196 | NA | 250 | NA | NA |
| Year of data collection | 2012 | 2019 – 2020 | 2018 – 2020 | 2016 | 2017 – 2020 | 2011 – 2012 |
| Initial TB tests used for diagnoses** | SSM, Xpert | Xpert, Xpert Ultra | Xpert, Xpert Ultra | Xpert | Xpert Ultra | SSM, Xpert |

**Table S1. Demographic and clinical characteristics of participants, by study.**

| | Pereira et al., 2020 | Hanrahan et al., 2015 | Mishra et al., 2020 | Luetkemeyer et al. 2016 | Bjerrum et al., 2020 | Agizew et al., 2019 |
|---|---|---|---|---|---|---|
| Study sites eligible for inclusion* | Brazil | South Africa | South Africa | Brazil, South Africa | Ghana | Botswana |
| Analytic population, n | 147 | 168 | 212 | 280 | 121 | 5838 |
| Individuals with treatment initiation, n (primary outcome) | 0 | 5 | 0 | 28 | 6 | 132 |
| Alternative outcome | 0 | 2 | 0 | 28 | 2 | 33 |
| Age category, n | | | | | | |
| 18 – 30 years | 24 | 25 | 68 | 54 | 33 | 2099 |
| 31 – 40 years | 12 | 66 | 48 | 92 | 42 | 2248 |
| 41 years and above | 111 | 77 | 96 | 134 | 46 | 1491 |
| Male, n | 76 | 120 | 113 | 133 | 38 | 1921 |
| Individuals with history of prior TB, n | 0 | 0 | 87 | 36 | 9 | 624 |
| Reported cough | | | | | | |
| None | 0 | 22 | 12 | 15 | 66 | 4793 |
| Yes | 147 | 146 | 200 | 265 | 55 | 1045 |
| Reported night sweats | | | | | | |
| None | 0 | 128 | 62 | 116 | 82 | 5321 |
| Yes | 147 | 40 | 150 | 164 | 39 | 517 |
| Reported fever | | | | | | |
| None | 49 | 132 | 186 | 134 | 60 | 5340 |
| Yes | 98 | 36 | 26 | 146 | 61 | 498 |
| HIV | | | | | | |
| Negative | 144 | 24 | 170 | 110 | 0 | 0 |
| Positive, not on ART | 3 | 38 | 42 | 170 | 38 | 5838 |
| Positive, on ART | 0 | 104 | 0 | 0 | 83 | 0 |
| Unknown | 0 | 2 | 0 | 0 | 0 | 0 |
| Individuals with abnormal chest X-ray result, n | NA | 10 | NA | NA | NA | NA |
| Year of data collection | 2018 – 2019 | 2011 – 2013 | 2016 – 2018 | 2012 – 2013 | 2013 – 2014 | 2012 – 2014 |
| Initial TB tests used for diagnoses** | Xpert Ultra | Xpert | SSM, Xpert, Xpert Ultra | SSM | Xpert | SSM. Xpert |

**Table S1 (continued). Demographic and clinical characteristics of participants, by study.**

* Data from countries with high income and low TB burdens were not considered for inclusion.
** Diagnostic tests performed solely for research purposes, which were not part of routine practice in the setting, were excluded.

| Country | Adjusted odds ratio of treatment initiation, compared to cross-country average (95% Credible Intervals) |
|---|---|
| Belarus | 0.56 (0.12, 2.04) |
| Botswana | 0.55 (0.26, 1.17) |
| Brazil | 0.33 (0.12, 0.83) |
| Ethiopia | 0.57 (0.16, 1.80) |
| Georgia | 0.48 (0.17, 1.17) |
| Ghana | 1.15 (0.42, 3.00) |
| India | 0.97 (0.46, 2.12) |
| Kenya | 2.14 (0.92, 5.09) |
| Papua New Guinea | 1.34 (0.42, 3.93) |
| Peru | 0.95 (0.31, 2.68) |
| South Africa | 0.36 (0.18, 0.72) |
| Uganda | 7.01 (3.43, 15.16) |
| Zimbabwe | 2.88 (1.32, 6.34) |

**Table S2. Odds ratios of TB treatment initiation following negative diagnostic test result: country random effects from primary analysis.**

| | Adjusted odds ratio of treatment initiation (95% Credible Intervals) |
|---|---|
| Age category | |
| 18 – 30 years | Ref |
| 31 – 40 years | 1.30 (0.53–3.26) |
| 41 years and above | 1.47 (0.71–3.25) |
| Sex | |
| Female | Ref |
| Male | 1.00 (0.56–1.83) |
| History of prior TB | |
| None | Ref |
| Yes | 0.64 (0.28–1.39) |
| Unknown | 0.50 (0.10–1.91) |
| Reported cough | |
| None | Ref |
| Yes | 1.26 (0.22–9.67) |
| Reported night sweats | |
| None | Ref |
| Yes | 1.14 (0.60–2.18) |
| Reported fever | |
| None | Ref |
| Yes | 0.68 (0.36–1.27) |
| HIV | |
| Negative | Ref |
| Positive, not on ART | 1.03 (0.17–5.44) |
| Positive, on ART | 1.28 (0.27–5.51) |
| Unknown | 0.58 (0.27–1.15) |
| Chest X-ray | |
| Normal | Ref |
| Abnormal | 6.89 (3.29–14.42) |
| Unknown | 0.69 (0.16–2.54) |
| Diagnostic test | |
| Sputum Smear | - |
| Xpert | Ref |
| Xpert Ultra | 0.77 (0.05–16.03) |
| Year | 0.86 (0.66–1.11) |

**Table S3. Odds ratios of TB treatment initiation following negative diagnostic test result: secondary analysis for datasets including chest x-ray results.**

| Study | Adjusted odds ratio of treatment initiation, compared to cross-study average (95% Credible Intervals) |
|---|---|
| Churchyard et al., 2015 | 0.53 (0.20, 1.50) |
| Penn-Nicholson et al., 2021 | 0.85 (0.27, 2.57) |
| Dorman et al., unpublished | 3.77 (1.30, 11.00) |
| Dorman et al., 2018 | 0.70 (0.28, 1.77) |
| Theron et al., unpublished | 1.14 (0.30, 4.05) |
| Mupfumi et al., 2014 | 4.97 (1.76, 15.56) |
| Pereira et al., 2020 | 0.27 (0.02, 1.67) |
| Hanrahan et al., 2015 | 1.30 (0.25, 9.72) |
| Mishra et al., 2020 | 0.10 (0.01, 0.48) |
| Luetkemeyer et al. 2016 | 1.18 (0.45, 3.21) |
| Bjerrum et al., 2020 | 1.68 (0.51, 5.79) |
| Agizew et al., 2019 | 0.87 (0.34, 2.33) |

**Table S4. Odds of TB treatment initiation following negative diagnostic test result: study random effects from alternative model specification.**

**Search terms for Embase (Elsevier, embase.com).**

Advanced Search:

Source: Embase subset (from 1974 to present)

Date: Publication years from 2010 - 2022

1) 'tuberculosis'/de OR 'lung tuberculosis'/de OR tuberculosis:ab,ti,kw

2) 'molecular diagnostics'/de OR 'molecular diagnosis'/de OR 'Mycobacterium tuberculosis test kit'/de OR 'polymerase chain reaction system'/de OR 'xpert'/de OR 'xpert ultra'/de OR 'xpert mtb rif ultra'/de OR 'sputum analysis'/de OR ('sputum smear'/de AND 'microscopy'/exp) OR 'sputum culture'/de OR genexpert:ab,ti,kw OR xpert:ab,ti,kw OR 'smear microscopy':ab,ti,kw OR 'sputum microscopy':ab,ti,kw

3) 'clinical trial'/de OR 'controlled clinical trial'/de OR 'randomized controlled trial'/exp OR 'diagnostic accuracy'/de OR 'diagnostic test accuracy study'/de OR 'evaluation study'/de OR 'clinical trial':ab,ti,kw OR random*:ab,ti,kw OR accuracy:ti,ab,kw OR evaluation:ab,ti,kw

1 AND 2 AND 3

NOT ('chapter'/it OR 'conference abstract'/it OR 'conference paper'/it OR 'conference review'/it OR 'editorial'/it OR 'review'/it)

**Search terms for MEDLINE/PubMed (National Library of Medicine, NCBI)**

("Tuberculosis"[Mesh:NoExp] OR "Tuberculosis, Pulmonary"[Mesh] OR tuberculosis[tiab]) AND ("Molecular Diagnostic Techniques"[Mesh:NoExp] OR ("Sputum"[Mesh] AND "Microscopy"[Mesh]) OR genexpert[tiab] OR xpert[tiab] OR smear microscopy[tiab] OR sputum microscopy[tiab]) AND ("randomized controlled trial"[pt] OR "controlled clinical trial"[pt] OR "random allocation"[mesh] OR "clinical trial"[pt] OR "evaluation study"[pt] OR "clinical trial"[tiab] OR random*[tiab] OR accuracy [tiab] OR evaluation[tiab]) AND 2010[pdat]: 2022[pdat].

**Hierarchical Bayesian logistic regression model.**

For the main analysis we fit the following regression model:

$$TREAT \sim Binomial\ (n = trials(1), p)$$

$$logit\ (p) = \beta_0 + \beta_{Age\_cat}Age_{cat} + \beta_{Sex}Sex + \beta_{TB_{hist}}TB_{hist} + \beta_{cough}Cough + \beta_{sweat}Sweat$$
$$+ \beta_{Fever}Fever + \beta_{HIV}HIV + \beta_{Diag_{test}}Diag_{test} + \beta_{year}Year + b_{country}$$

Where:

- $TREAT$ is the binary outcome variable indicating whether or not an individual initiated TB treatment.
- $p$ is the probability of receiving treatment.
- $Age_{cat}$ , $Sex, TB_{hist}, Cough, Sweat, Fever, HIV, Diag_{test}$, and $Year$ are exposure variables as defined in Table 2.
- $b_{country}$ represents the country random effect term.

All analyses were performed using the “brms” package (v.2.19.0) in R (v.4.2.3). We adopted the default prior distributions available in this package, with all main effects given prior student-*t* distributions with 3 degrees of freedom and a scale parameter of 10, and the random effects standard deviation given a half student-*t* prior with 3 degrees of freedom. The approach uses an extension of Hamiltonian Monte Carlo to sample from the posterior distribution of the regression model parameters (24–26). During the sampling process, 4,000 posterior samples were generated and convergence was assessed using Gelman-Rubin statistic (i.e., potential scale reduction factor (PSRF)). PSRF values for all parameters were observed to be 1.00, confirming convergence (39). The reported coefficient estimates (aOR) in **Table 2** represent the posterior means for each parameter and equal-tailed 95% credible intervals (95% CI). Secondary analyses and alternative model specifications were performed using a similar approach.

**Description of individual studies included in analysis.**

The XTEND study was a pragmatic cluster-randomized trial conducted in South Africa assessing 6-month mortality of clinic attendees randomly allocated testing with Xpert MTB/RIF or sputum smear microscopy (6). Penn-Nicholson et al. conducted a prospective multi-center diagnostic accuracy study conducted in four countries (Peru, India, Ethiopia and Papua New Guinea) to assess the performance of the Truenat TB assays compared to Xpert MTB/RIF (40). Dorman et al. conducted a prospective multi-center diagnostic accuracy study in eight countries (South Africa, India, Georgia, and Belarus) assessing sensitivity and specificity of Xpert Ultra compared to Xpert (32). Dorman et al. subsequently conducted the Ultra 2 study in three countries (South Africa, Uganda, Kenya), incorporating the same study procedures as used in the Ultra study (32), with improvement in stability of the initial Ultra assay (Dorman et al., unpublished). Theron and the study team conducted a prospective diagnostic accuracy study in South Africa focusing on people living with HIV (PLHIV) attending clinics to start antiretroviral therapy (ART). The preliminary findings were published in the Union Conference. Mupfumi et al. conducted a pragmatic randomized control trial in Zimbabwe, examining the impact of Xpert on ART-associated TB and patient outcomes (10). Pereira et al assessed the diagnostic accuracy of Xpert Ultra in Brazil (41). Hanrahan et al. conducted a prospective cohort study in South Africa to investigate the effects of placement of Xpert at the point of care (POC) (42). Mishra et al. conducted a two-cohort diagnostic accuracy study in South Africa (43). Luetkemeyer et al. conducted a longitudinal multicenter study in the US, Brazil and South Africa to evaluate the Xpert assay (44). Bjerrum et al. conducted a diagnostic accuracy study of LAM studies among PLHIV in Ghana (45). Lastly, Agizew et al. conducted a stepped-wedge cluster randomized trial comparing TB treatment outcomes of SSM and Xpert among PLHIV in Botswana (46).
